# Development and validation of a clinical risk score to predict SARS-CoV-2 infection in emergency department patients: The CCEDRRN COVID-19 Infection Score (CCIS)

**DOI:** 10.1101/2021.07.15.21260590

**Authors:** Andrew D McRae, Corinne M Hohl, Rhonda J Rosychuk, Shabnam Vatanpour, Gelareh Ghaderi, Patrick M Archambault, Steven C Brooks, Ivy Cheng, Philip Davis, Jake Hayward, Eddy S Lang, Robert Ohle, Brian H. Rowe, Michelle Welsford, Krishan Yadav, Laurie J Morrison, Jeffrey J. Perry, Canadian COVID-19 Emergency Department Rapid Response Network (CCEDRRN) investigators for the Network of Canadian Emergency Researchers and the Canadian Critical Care Trials Group

**Author notes:** co-first authors (Andrew D McRae, Corinne M Hohl). co-senior author (Laurie J Morrison, Jeffrey J. Perry). Corresponding Author: Andrew D. McRae Department of Emergency Medicine Rm C231 Foothills Medical Centre 1403 29 St NW. Calgary, AB, Canada. T2N 2T9.

## Abstract

**Objectives:** To develop and validate a clinical risk score that can accurately quantify an emergency department patient’s probability of SARS-CoV-2 infection without the need for laboratory testing

**Design:** Cohort study of participants in the Canadian COVID-19 Emergency Department Rapid Response Network (CCEDRRN) registry. Regression models were fitted to predict a positive SARS-CoV-2 test result using clinical and demographic predictors, as well as an indicator of local SARS-CoV-2 incidence.

**Setting:** 32 emergency departments in eight Canadian provinces

**Participants:** 27,665 consecutively-enrolled patients who were tested for SARS-CoV-2 in participating emergency departments between March 1-October 30,2020

**Main outcome measures:** Positive SARS-CoV-2 nucleic acid test result within 14 days of an index emergency department encounter for suspected COVID-19 disease

**Results:** We derived a 10-item CCEDRRN COVID-19 Infection Score using data from 21,743 patients. This score included variables from history and physical examination, and an indicator of local disease incidence. The score had a c-statistic of 0.838 with excellent calibration. We externally validated the rule in 5,295 patients. The score maintained excellent discrimination and calibration, and had superior performance compared to another previously published risk score. Score cutoffs were identified that can rule-in or rule-out SARS-CoV-2 infection without the need for nucleic acid testing with 97.4 % sensitivity (95% CI 96.4–98..3) and 95.9% specificity (95% CI 95.5-96.0).

**Conclusions:** The CCEDRRN COVID-19 Infection Score uses clinical characteristics and publicly available indicators of disease incidence to quantify a patient’s probability of SARS-CoV-2 infection. The score can identify patients at sufficiently high risk of SARS-CoV-2 infection to warrant isolation and empiric therapy prior to test confirmation, while also identifying patients at sufficiently low risk of infection that they may not need testing.

**Trial registration:** CCEDRRN is registered at clinicaltrials.gov (NCT04702945).

**Funding:** The network is funded by the Canadian Institutes of Health Research (447679), BC Academic Health Science Network Society, BioTalent Canada, Genome BC (COV024; VAC007), Ontario Ministry of Colleges and Universities (C-655-2129), the Saskatchewan Health Research Foundation (5357) and the Fondation CHU de Québec (Octroi #4007). These organizations are not-for-profit, and had no role in study conduct, analysis, or manuscript preparation.

## Introduction

To date, the World Health Organization has reported 170 million diagnosed cases of coronavirus 2019 disease (COVID-19) with 3.5 million fatalities.^1^ Despite the availability of vaccines to prevent COVID-19, incomplete population-level immunization and the emergence of variants means that hospitals around the world need to continue to identify and isolate patients with suspected COVID-19 from the time they arrive in the emergency department until their SARS-CoV-2 test results are available. In acutely ill patients, clinicians may need to initiate empiric therapy immediately. A quantitative risk score that can accurately predict the probability of a positive SARS-CoV-2 test result would guide initial isolation and empiric therapy prior to nucleic acid amplification test (NAAT) test result availability, while identifying patients with sufficiently low probability of COVID-19 who may not require testing or isolation.

Many risk prediction tools have been developed to predict the probability of SARS-CoV-2 infection.^2–14^ A living systematic review of these models concluded that most were generated using poor methodological approaches and none were ready for widespread use.^2^ Most published risk prediction tools included early laboratory or imaging findings, thus precluding their utility to guide immediate isolation and clinical decisions at the time of first clinical contact. Other risk prediction tools using machine learning included laboratory and imaging results and can only be implemented in hospitals using electronic health records with integrated decision support. None of these models accounted for the prevalence of COVID-19 disease in the local population, which is an important risk predictor, and most only included patients from the early stages of the pandemic.^2^

The objective of this study is to develop a clinical risk score to predict the probability of a positive SARS-CoV-2 nucleic acid test in a large, generalizable population of emergency department patients using only clinical characteristics and indicators of local SARS-CoV-2 incidence. This risk score is intended to guide SARS-CoV-2 testing, isolation, and empiric therapy decisions without relying on other laboratory testing or diagnostic imaging. This score could be invaluable in settings that may not have access to adequate resources for timely SARS-CoV-2 testing.

## Methods

This analysis uses data from the Canadian COVID-19 Emergency Department Rapid Response Network (CCEDRRN, pronounced “SED-rin”). CCEDRRN is an ongoing multicenter, pan-Canadian registry that has been enrolling consecutive emergency department patients with suspected COVID-19 disease in hospitals in eight of ten Canadian provinces since March 1, 2020.^15^ Information on the network, including detailed methods and participating sites, is available elsewhere.^15^ This study follows the methodological and reporting recommendations outlined in the Transparency in reporting of a multivariable prediction model for individual diagnosis and prognosis (TRIPOD) criteria.^16^ The CCEDRRN network protocol was approved by the research ethics boards of all participating institutions with a waiver of informed consent for data collection and linkage.

The CCEDRRN data collection form includes prespecified demographic and social variables, vital signs, symptoms, and comorbid conditions (derived from the International Severe Acute Respiratory and Emerging Infection Consortium (ISARIC) reporting form),^17, 18^ exposure risk variables, hospital laboratory and diagnostic imaging test results, SARS-CoV-2 NAAT results, and patient outcomes. Data were abstracted at each site using electronic medical record extraction where available as well as manual review of either electronic or paper charts (depending on site-specific documentation practices) by trained research assistants who were blinded to the potential predictor variables at the time of data collection. Reliability of health record data abstraction was evaluated by comparison with prospective data collection in a sample of patients and found to be reliable.^15^

Each consecutive, eligible patient enrolled in the registry was assigned a CCEDRRN unique identifier. Trained research assistants entered anonymized participant data into a REDCap database (Version 10.9.4; Vanderbilt University, Nashville, Tennessee, USA). Regular data quality checks including verification of extreme or outlying values were performed by each participating site, coordinated by the CCEDRRN coordinating center.

### Participants

We included data from consecutive patients presenting to 32 CCEDRRN sites that collected data on all patients tested for SARS-CoV-2. We included consecutive eligible patients aged 18 and older who had a biological sample (swab, endotracheal aspirate, bronchoalveolar lavage) specimen collected for NAAT on their index emergency department visit or, if admitted, within 24h of emergency department arrival. For patients with multiple emergency department encounters involving COVID-19 testing, we only used the first encounter in this analysis.

We excluded patients who had a positive SARS-CoV-2 NAAT within 14 days prior to their emergency department visit, patients with cardiac arrest prior to emergency department arrival, and those with missing outcome data.

### Predictors

Candidate predictors were chosen based on clinical consensus and availability within the CCEDRRN registry. Predictors included known risk factors for SARS-CoV-2 infection, including work as a healthcare provider, institutional living (i.e., long term care, prison), close personal or household contacts with SARS-CoV-2 infection; symptoms including cough, anosmia or dysgeusia, fever, myalgias and vital signs on emergency department arrival. The full list of candidate variables, and their definitions are available in the supplementary appendix (Appendix Table 2).

In addition to these clinical variables, the seven-day average incident COVID-19 case count was calculated for the health region of each participating site using publicly available epidemiological data.^19^ For each calendar day within each health region represented in the study, we calculated the average daily incident rate of new infections per 100,000 population over the preceding seven days. This seven-day average incidence was assigned to each patient based on the date of their index emergency department encounter and the health region of the forward sortation area of their postal code of residence. For patients with no fixed address, we allocated them to the health region of the hospital in which they were tested. As publicly available incident COVID-19 case data were not available for the early pandemic, we imputed values for the first five weeks of the pandemic by modeling the reported COVID-19 cases that had accumulated in every health region over time using linear interpolation (0.1% missing).

### Outcome

The primary outcome of this analysis was the diagnosis of SARS-CoV-2 infection using a criterion standard of a positive NAAT at the time of index emergency department visit or within 14 days after the index encounter.

### Sample size and precision

The 46 candidate predictors had 52 degrees of freedom and with an expected SARS-CoV-2 infection rate of 5%, a sample size of 1040 was sufficient for the derivation cohort based on an anticipated event rate of less than 20% and a requirement for 20 outcomes per degree of freedom.^20^ Over 21,000 patients were available for the derivation cohort at the time of analysis, providing more than sufficient data for reliable prediction modeling.

### Model development and validation

We randomly assigned study sites to the derivation and validation cohorts with the goal of assigning 75% of eligible patients and outcome events to the derivation cohort and 25% to the validation cohort. Thus, the derivation and validation cohorts are geographically distinct. Within the derivation cohort, candidate predictors were examined for co-linearity and missing or extreme values. In the presence of co-linearity, one predictor was dropped from the set of candidate predictors. Five multiple imputations were used for continuous variables with missing data. Patients with missing data for categorical variables were assumed to have the reference value for that categorical variable. The initial logistic regression model considered all candidate predictors, with continuous predictors fit with restricted cubic splines with three knots. The strengths of associations between predictors and outcome were assessed using an analysis of variance (ANOVA) plot to inform the degrees of freedom to allocate to each predictor. The model was fit again with these changes. A fast step-down procedure reduced the model to key predictors based on an Akaike’s information criterion stopping rule with a threshold of 120 to enable a model with a relatively small number of predictors that would be clinically easy to use. Internal bootstrap validation with 1,000 bootstrap samples was conducted to provide an optimism-corrected C-statistic. Continuous predictors were categorized based on the relationship between the spline function and outcome.

We then developed the points-based CCEDRRN COVID-19 Infection Score (CCIS) using a nomogram to assign integer point values for each variable included in the derived model. Discrimination of the score was evaluated using the C-statistic. Calibration was evaluated using calibration curves and comparison of observed and expected outcomes. Diagnostic performance was evaluated using sensitivity and specificity, predictive values, and likelihood ratios at different point thresholds.

We then evaluated the discrimination, calibration, and performance characteristics of the CCIS in an external validation cohort of patients from geographically distinct study sites who were not part of the derivation cohort.

### Validation of previously published models

We used our combined (derivation and validation) study cohort to externally validate the COvid Rule out Criteria (CORC) score developed by Kline et al (with race and ethnicity variables removed).^3^ We compared measures of discrimination and calibration, along with sensitivity and specificity of risk score values for the CCIS and CORC (with race and ethnicity variables removed). We split each score into categories of low, moderate, and high-risk for SARS-CoV-2 infection. Low risk was defined as a score having a sensitivity for ruling-out infection of 95% or higher. High-risk was defined as a score having a specificity for ruling in infection of 95% or higher. We compared the performance of the two scores by calculating net reclassification improvement across low, moderate, and high-risk categories.^21, 22^

All analyses were performed in R ^23^ using the rms package.^24^

### Patient involvement

The CCEDRRN governance structure includes patient representatives on the Executive Committee, Scientific Steering Committee, Protocol Review and Publications Committee, Data Access and Monitoring Committee and Knowledge Translation Committee. The network also has a Patient Engagement Committee composed of patient partners from across Canada. Patient partners provided input into study design and selection of outcomes for all CCEDRRN analyses, and provide advice on knowledge sharing and translation strategies.

## Results

This analysis is based on 27,665 consecutively enrolled patients from 32 participating emergency departments (Figure 1, Appendix Table 1). Sites and enrolment periods contributing patient data are shown in the supplementary appendix. Of the included patients, 1,677 (4.2%) had a positive SARS-CoV-2 NAAT result.

**Figure 1.**
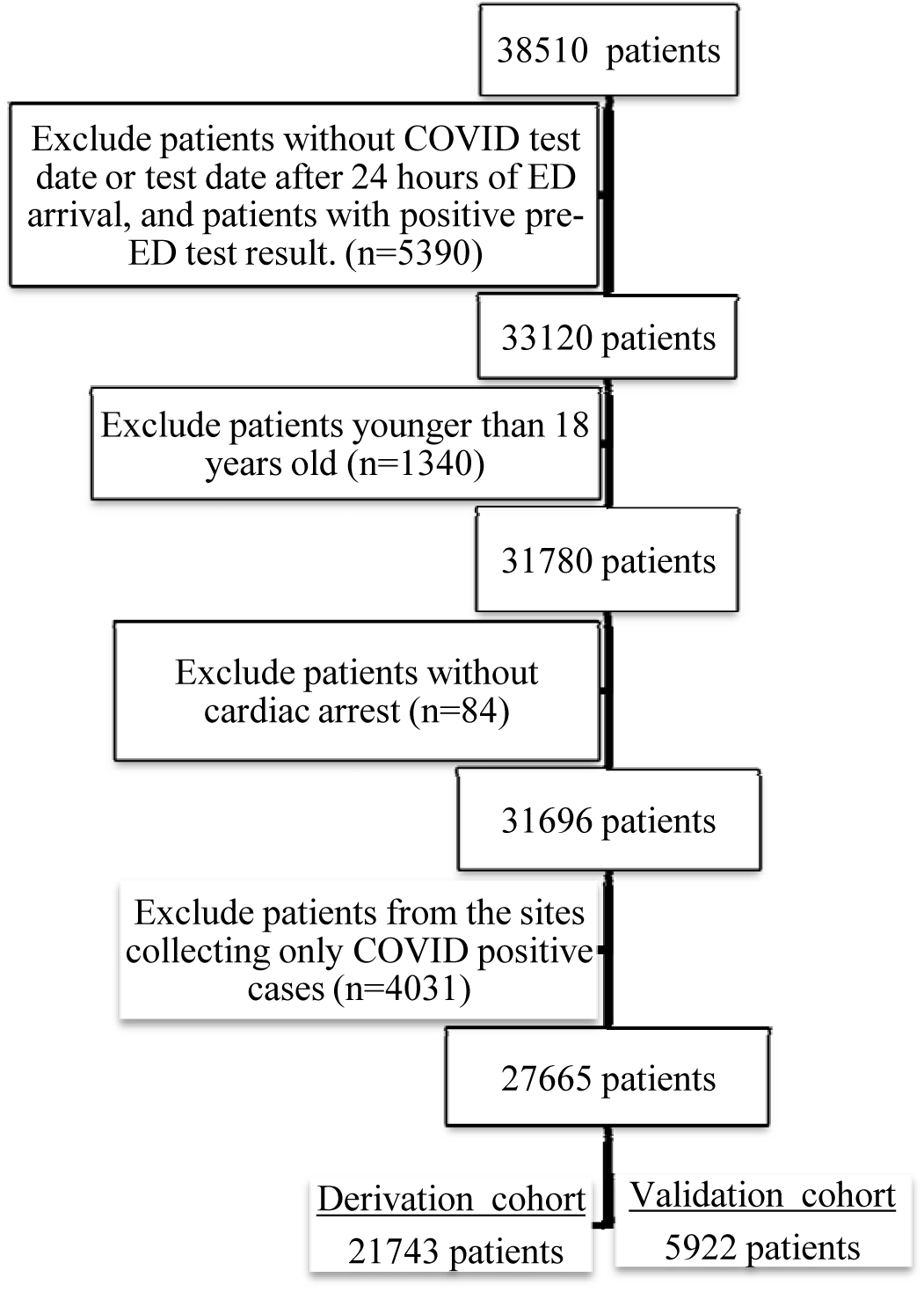
Flow Diagram of Patients through the study (based on PRPC template)

The study cohort was subdivided into a derivation cohort (21,743 patients from 16 sites, 940 (4.3%) SARS-CoV-2 positive) and a separate external validation cohort (5,922 patients from 16 different sites, 227 (3.8%) SARS-CoV-2 positive). Demographic and clinical characteristics of the derivation and validation cohorts are shown in Table 1. No continuous variable requiring multiple imputation had more than 3.4% missingness (Appendix Table 2).

**Table 1.** Characteristics and selected outcomes of enrolled patients·

|  | **Derivation**<br>**(n=21743)** | **Validation**<br>**(n=5922)** |
| --- | --- | --- |
| **Age in years, median (IQR)** | 57 (38, 73) | 56 (37, 73) |
| **Female (%)** | 10992 (50.5) | 3085 (52.1) |
| **Arrival From, n (%)** |  |  |
| Home | 19879 (91.4) | 5429 (91.7) |
| Long-term care/Rehabilitation facility/Corrections facility | 1000 (4.6) | 262 (4.4) |
| No fixed address/ Shelter/ Single room occupancy | 574 (2.6) | 201 (3.4) |
| Inter-hospital transfer | 290 (1.3) | 30 (0.5) |
| **Risk for Infection, n (%)** |  |  |
| Healthcare worker | 505 (2.3) | 567 (9.6) |
| Household/caregiver contact | 566 (2.6) | 161 (2.7) |
| Institutional exposure (e.g., LTC, prison) | 1354 (6.2) | 213 (3.6) |
| Microbiology lab | 4 (0.0) | 8 (0.1) |
| Travel | 924 (4.2) | 344 (5.8) |
| Other | 1320 (6.1) | 449 (7.6) |
| Unknown | 5415 (24.9) | 1856 (31.3) |
| No documented risk for infection | 10028 (46.1) | 1075 (18.1) |
| **Arrival Vital Signs, median (IQR)** |  |  |
| Body temperature | 36.7 (36.3, 37.1) | 36.8 (36.5, 37.1) |
| Heart rate | 91 (79, 107) | 90 (78, 105) |
| Oxygen saturation | 97 (95, 98) | 97 (95, 99) |
| Respiratory rate | 18 (18, 20) | 18 (16, 20) |
| Systolic blood pressure | 133 (118, 150) | 136 (120, 149) |
| **Common Comorbid Conditions, n (%)** |  |  |
| Active malignant neoplasm (cancer) | 1678 (7.7) | 333 (5.6) |
| Asthma | 1699 (7.8) | 468 (7.9) |
| Atrial fibrillation | 1598 (7.3) | 402 (6.8) |
| Chronic kidney disease | 1214 (5.6) | 321 (5.4) |
| Chronic lung disease (not asthma/pulmonary fibrosis) | 1729 (8) | 583 (9.8) |
| Chronic neurological disorder (not dementia; e.g., stroke/TIA, seizure disorder) | 1310 (6) | 400 (6.8) |
| Congestive heart failure | 1450 (6.7) | 368 (6.2) |
| Coronary artery disease | 1591 (7.3) | 449 (7.6) |
| Dementia | 734 (3.4) | 188 (3.2) |
| Diabetes | 2583 (11.9) | 916 (15.5) |
| Dialysis | 198 (0.9) | 28 (0.5) |
| Dyslipidemia | 2375 (10.9) | 543 (9.2) |
| Hypertension | 6320 (29.1) | 1697 (28.6) |
| Hypothyroidism | 1344 (6.2) | 397 (6.7) |
| Mild liver disease | 280 (1·3) | 90 (1·5) |
| Moderate/severe liver disease | 245 (1·1) | 88 (1·5) |
| Obesity (clinical impression) | 284 (1·3) | 108 (1·8) |
| Organ transplant | 128 (0·6) | 19 (0·3) |
| Rheumatologic disorder | 1122 (5·2) | 258 (4·4) |
| Other | 10075 (46·3) | 2174 (36·7) |
| Past malignant neoplasm (cancer) | 936 (4·3) | 256 (4·3) |
| Psychiatric condition/Mental health diagnosis | 2967 (13·6) | 831 (14) |
| Pulmonary fibrosis | 80 (0·4) | 26 (0·4) |
| **Symptoms Reported, n(%)** |  |  |
| Abdominal pain | 2725 (12·5) | 540 (9·1) |
| Altered consciousness/confusion | 1456 (6·7) | 322 (5·4) |
| Bleeding (hemorrhage) | 330 (1·5) | 22 (0·4) |
| Chest pain (includes discomfort or tightness) | 4242 (19·5) | 974 (16·4) |
| Chills | 2045 (9·4) | 594 (10) |
| Conjunctivitis | 49 (0·2) | 26 (0·4) |
| Cough | 7724 (35·5) | 2663 (44·9) |
| Diarrhea | 2140 (9·8) | 526 (8·9) |
| Dizziness/Vertigo | 1521 (7) | 300 (5·1) |
| Dysgeusia/anosmia | 140 (0·6) | 33 (0·6) |
| Ear pain | 144 (0·7) | 30 (0·5) |
| Fatigue/malaise | 3361 (15·5) | 924 (15·6) |
| Fever | 5055 (23·2) | 1580 (26·7) |
| Headache | 2144 (9·9) | 624 (10·5) |
| Hemoptysis (bloody sputum) | 298 (1·4) | 66 (1·1) |
| Joint pain (arthralgia) | 296 (1·4) | 82 (1·4) |
| Lower chest wall indrawing | 10 (0) | 7 (0·1) |
| Lymphadenopathy | 67 (0·3) | 21 (0·4) |
| Muscle aches (myalgia) | 1575 (7·2) | 517 (8·7) |
| Nausea/vomiting | 4219 (19·4) | 935 (15·8) |
| No recorded symptoms | 2113 (9·7) | 431 (7·3) |
| Runny nose (rhinorrhea) | 1061 (4·9) | 501 (8·5) |
| Seizures | 205 (0·9) | 42 (0·7) |
| Shortness of breath (dyspnea) | 8537 (39·3) | 2383 (40·2) |
| Skin rash | 241 (1·1) | 38 (0·6) |
| Skin ulcers | 27 (0·1) | <5 |
| Sore throat | 3024 (13·9) | 985 (16·6) |
| Sputum production | 1507 (6·9) | 401 (6·8) |
| Wheezing | 582 (2·7) | 130 (2·2) |
| **Tobacco Use, n (%)** | 1852 (8·5) | 616 (10·4) |
| **Illicit Substance Use, n (%)** | 1219 (5·6) | 353 (6·0) |
| **Oxygen Required in ED, n (%)** | 1919 (8·8) | 627 (10·6) |
| **Hospital Admission, n (%)** | 9913 (45·6) | 2446 (41·3) |
| **In-hospital Death, n (%)** | 753 (3·5) | 213 (3·6) |
| **7-day average incident COVID-19 cases, median (IQR)** | 1·3 (0·7, 3·2) | 0·96 (0·5, 1·3) |
| **SARS-CoV-2 Positive, n (%)** | 940 (4·3) | 227 (3·8) |

IQR=interquartile range; LTC=long-term care; TIA= transient ischemic attack; ED=emergency department

In the derivation cohort, we derived a 10-variable model to predict the probability of a patient having a positive SARS-CoV-2 NAAT. The regression coefficients and odds ratios for each variable in the model are shown in Table 2. The C-statistic for the derived model was 0.851 with excellent calibration.

**Table 2.** Adjusted associations between model predictor variables and SARS-CoV-2 nucleic acid test results

| **Variable/Score Component** | **Regression Coefficient (SE)** | **Adjusted Odds Ratio (95% CI)** | **Score Value** |
| --- | --- | --- | --- |
| **7-day average incident COVID-19 cases** |  |  |  |
| 0 – 2 daily cases per 100,000 population | - | - | 0 |
| 2 to 7.99 daily cases per 100,000 population | 1.22 (0.09) | 3.38 (2.85– 4.00) | 1 |
| ≥8 daily cases per 100,000 population | 2.21 (0.10) | 9.09 (7.53– 10.97) | 2 |
| **Institutional exposure (e.g. LTC, prison) or Travel from country with known cases within 14 days** | 0.88 (0.09) | 2.40 (2.01–2.87) | 1 |
| **Healthcare worker/Microbiology lab** | 1.10 (0.16) | 3.02 (2.22–4.10) | 1 |
| **Household/caregiver contact** | 1.83 (0.12) | 6.25 (4.92–7.93) | 2 |
| **Temperature** |  |  |  |
| <36 and no self-reported fever | -0.75 (0.3) | 0.47 (0.28–0.80) | -1 |
| 36 – 37.4 and no self-reported fever | - | - | 0 |
| ≥37.5 or self-reported fever | 1.21 (0.08) | 3.36 (2.88–3.91) | 1 |
| **Supplemental oxygen delivered in the ED** | 0.98 (0.1) | 2.66 (2.18–3.24) | 1 |
| **Cough** | 0.85 (0.08) | 2.33 (2.01–2.71) | 1 |
| **Dysgeusia/Anosmia** | 2.03 (0.24) | 7.60 (4.76–12.15) | 2 |
| **Muscle aches (Myalgia)** | 0.7 (0.11) | 2.02 (1.64–2.48) | 1 |
| **Current tobacco user** | -1.13 (0.21) | 0.32 (0.21–0.49) | -1 |

LTC: Long-term care; ED: Emergency Department

We created a points-based CCEDRRN COVID-19 Infection Score (CCIS) using rounded regression coefficients with a range of negative two to nine points (Table 2). The C-statistic of the CCIS in the derivation cohort was 0.838 (0.824–0.852) with excellent calibration (Figure 2). A score of zero or less ruled out a positive SARS-CoV-2 test result in 5,996/21,743 patients (27.6%) with a sensitivity of 96.6% (95% CI 95.2–97.7). A score of four or more was observed in 1,338/21,743 patients (6.2%) and had a specificity of 95.6 (95% CI 95.3–95.8) for predicting a positive SARS-CoV-2 test result (Appendix Table 3).

**Figure 2.**
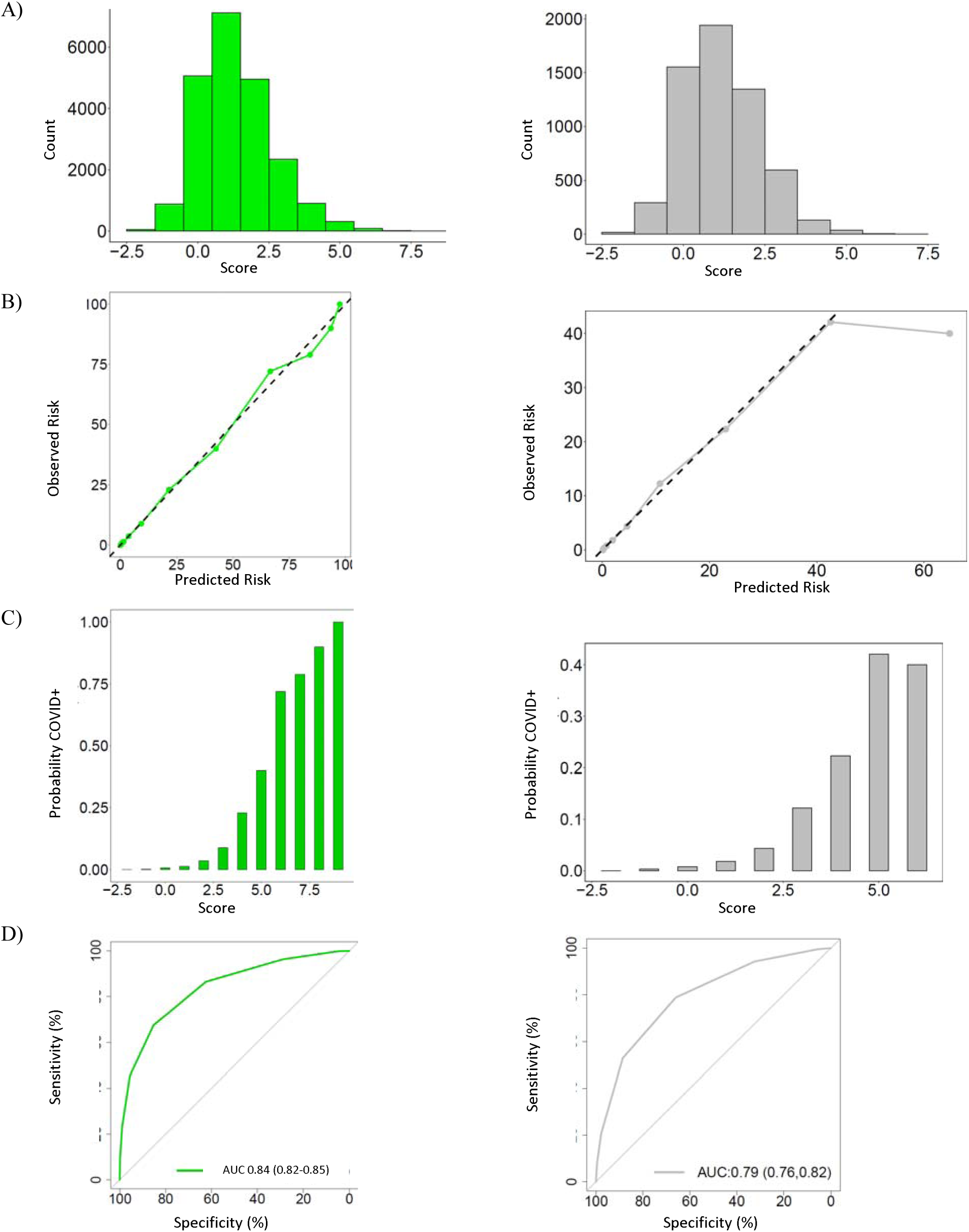
Distribution and performance of the CCEDRRN COVID Infection Score in the derivation cohort (left panel) and validation cohorts (right panel): A) distribution of the score, B) observed in-hospital mortality across the range of the score, C) predicted versus observed probability of in-hospital mortality, and D) receiver operating characteristic curve with area under the curve (AUC) and associated 95% confidence interval.

We then quantified the performance of the CCIS in our external validation cohort. In this cohort, the C-statistic for the points-based risk score was 0.792 (Figure 2). A score of zero or less ruled out a positive SARS-CoV-2 test result in 1,863/5,925 patients (31.4%) with a sensitivity of 94.3% (95% CI 90.4–96.9). A score of four or more was observed in 174/5,925 patients (2.9%) and had a specificity of 97.8 (95% CI 97.4–98.1) for predicting a positive SARS-CoV-2 test result (Table 3).

**Table 3.**
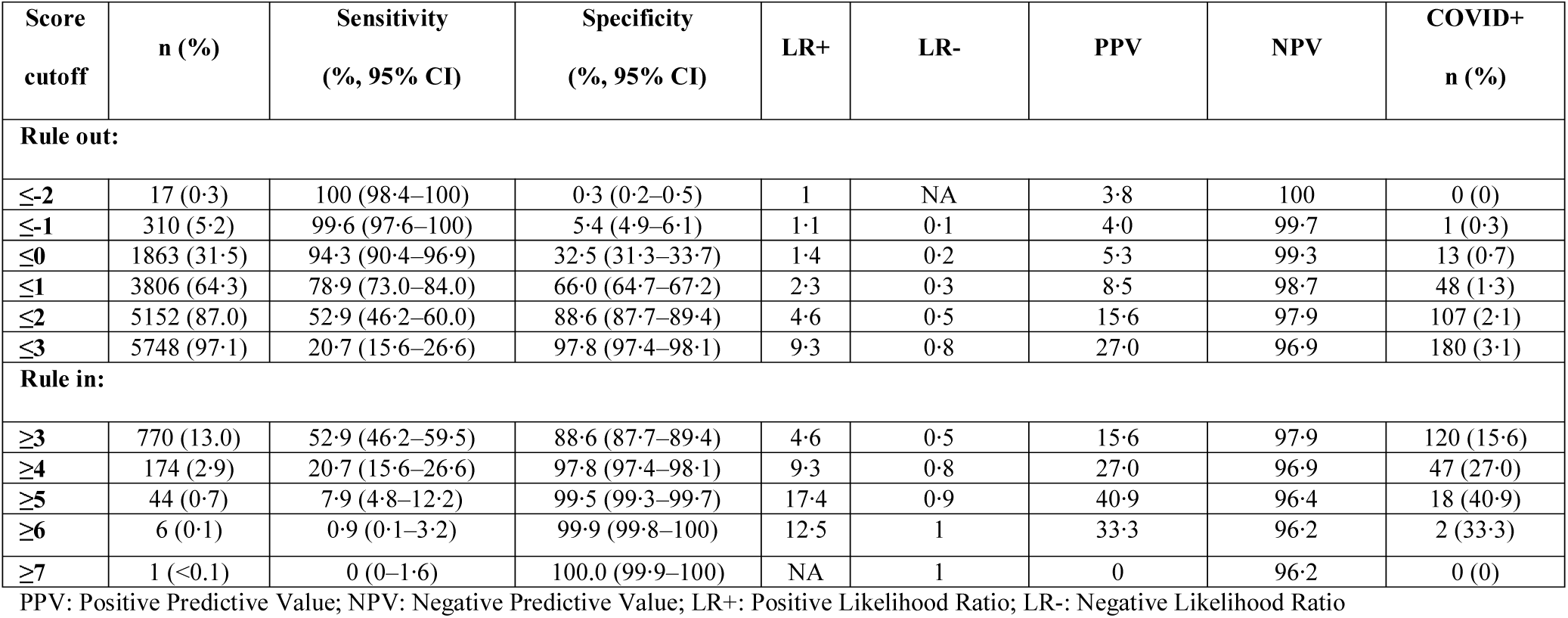
Performance metrics for the CCEDRRN COVID-19 Infection Score for ruling in or ruling out SARS-CoV-2 infection at different score cut-off values in the validation cohort

In a combined cohort of patients (derivation and validation combined), we compared the discrimination and diagnostic performance of the CCIS to the CORC score. The CCIS had a C-statistic of 0.837 compared to 0.750 for the CORC score (with race/ethnicity variables removed) (Appendix Figure 1). A CCIS of zero or less ruled out SARS-CoV-2 infection in 28.4% of patients with a sensitivity of 96.1% (Appendix Table 4) whereas a CORC score of negative one or less ruled out SARS-CoV 2 infection in 9.9% of patients with 97.4% (Appendix Table 5) sensitivity. Compared to the CORC score (with race/ethnicity variables removed), the CCIS showed substantial net reclassification improvement (NRI=0.310, Appendix Table 6).

## Discussion

We have derived and validated a simple clinical risk score, the CCEDRRN COVID-19 Infection Score (CCIS), to predict an emergency department patient’s probability of a positive SARS-CoV-2 NAAT. It utilizes only clinical variables available at the patient’s bedside, along with a common publicly available measure of community COVID-19 incidence. In this study population, the score ruled out SARS-CoV-2 infection with 96.1% sensitivity in almost one-third of patients. It also identified patients at high risk of infection with over 95% specificity.

The CCIS has several important clinical applications. The ability to differentiate patients with high or low probability of COVID-19 disease could guide safe and effective patient isolation or cohorting from the time of hospital arrival, prior to the availability of SARS-CoV-2 test results. Identification of patients with extremely low risk of SARS-CoV-2 infection may even allow safe omission of testing, which will minimize testing resource utilization in settings with limited testing capacity. Identifying patients with a high probability of SARS-CoV-2 infection can help prioritize use of rapid antigen testing and initiation of effective empiric therapy in critically ill patients prior to availability of NAAT results. By presenting risk estimates and sensitivity for all risk score values, we allow end-users to choose cut-offs for ruling-in and ruling-out SARS-CoV-2 infection that make sense for their setting and application.

Several other risk prediction instruments have been developed to predict positive COVID-19 test results in undifferentiated patients. These tools were developed in studies with substantial methodological limitations and incorporate variables not immediately available at the time of a patient’s hospital arrival, so are not useful to guide early isolation, testing and treatment decisions.^2^ None of these risk prediction tools considered the prevalence of disease in the population. Prevalence can substantially change the approach to testing and cohorting, and this will become increasingly important as prevalence rates drop and selective rather than liberal testing may be more appropriate.

United States-based investigators recently reported the development of the CORC score using only clinical variables. The CORC score contains several similar variables to the CCIS. However, the CORC score was derived in a non-consecutive sample of patients which had a much higher incidence of disease than our study cohort and may be vulnerable to selection bias. The CORC score also included race and ethnicity as predictor variables. This inclusion of race and ethnicity variables limits the generalizability of the CORC score beyond the urban American population in which it was developed, as it does not reflect the international diversity of ethnic backgrounds. Moreover, it is unlikely race or ethnicity represents a biologic risk. The association between race and ethnicity and SARS-CoV-2 infection in the CORC score likely reflects other sociodemographic and geographic predictors of the risk of COVID-19 infection in the American population.^25^ The CCIS was derived in consecutive patients with a suspected SARS-CoV-2 infection presenting to participating emergency departments, limiting potential for selection bias, and uses the seven-day average local incidence as an estimate of population risk. We believe this approach is more generalizable across populations and better reflects individual patients’ pre-test probability of SARS-CoV-2 infection.^25^

### Strengths and Limitations

The cohorts used to derive and validate the rule included comprehensive data on consecutive eligible patients from a large, geographically distributed network of Canadian urban, regional, and rural emergency departments. Strict data quality protocols and data cleaning protocols ensured the reliability of collected data.

In addition to clinical variables, we also included the average daily incidence of SARS-CoV-2 infections in a patient’s health region, which is an essential predictor of the probability of a patient’s risk of COVID infection. This information is publicly reported in many health jurisdictions and particularly in high- and low-prevalence regions. This information remains constant over long periods of time so it can easily be integrated into risk prediction for an individual patient. In practical application of this risk score, patients in areas with high disease burden will automatically score two points, meaning that few patients in these settings will be classified as low risk. Therefore, symptomatic patients would all warrant testing. This underscores the need for liberal isolation and testing practices in settings with high rates of community SARS-CoV-2 transmission.

This study has some limitations. Some missing data required either multiple imputation or classification of missing categorical variables as being absent. The overall missingness of data in this registry is very low. ^15^ Although the data collection for the CCEDRRN registry relies on abstraction from health records, this approach has been shown to be reliable in our study sites when compared to prospective data collection.^15^

The clinical variables in the model are not likely to be sensitive to changes in geographical changes in SARS-CoV-2 epidemiology. The variable of travel from a country with high incidence may become less informative as the pandemic has spread globally and “hot spots” change. However, high-prevalence areas may change over time, meaning that the risk factor of travel from a region with a high prevalence is likely to still be informative.

This risk score was developed using data from patients enrolled in the first nine months of the pandemic when rates of influenza were low. As such, the score may need to be re-validated and refined in the future to reflect the influence of influenza, the emergence of variant strains of SARS-CoV-2, and widespread population immunization on patients’ risk of infection.

### Conclusion

We derived and successfully validated the CCEDRRN COVID-19 Infection Score to accurately predict the probability of SARS-CoV-2 nucleic acid test results in emergency department patients. The CCIS uses clinical variables, accounts for the incidence of SARS-CoV-2 in the community and is ready for immediate clinical use. This score has potential utility to guide early decisions around SARS-CoV-2 test utilization, patient isolation, and empiric therapy for patients solely based on clinical assessment.

## Data Availability

The CCEDRRN network accepts applications for access to data by external investigators, prioritizing data requests by network Members.

https://canadiancovid19ednetwork.org/data-access/

## Contributors

CMH, ADM, LJM, RJR, and JJP conceived the study, with input on the design and selection of variables from the other contributors. CMH, LJM, PA, SCB, PD obtained funding on behalf of the CCEDRRN investigators. CMH, ADM, PA, SCB, IC, PD, JH, BHR, RO, MW, and KY facilitated data collection along with other members of the CCEDRRN and can verify the underlying data. RJR and JJP developed the analytic plan. SV performed the analysis, with assistance from GG and RJR, including accessing and verification of underlying data. All contributors provided input on interpretation of findings. ADM, CMH, and RJR drafted the manuscript with additional input from all contributors.

## Acknowledgment

We gratefully acknowledge the assistance of Ms. Amber Cragg in the preparation of this manuscript. We thank the UBC clinical coordinating centre staff, the UBC legal, ethics, privacy and contract staff and the research staff at each of the participating institutions in the network outlined in the attached Supplement. The network would not exist today without the dedication of these professionals.

Thank you to all our patient partners who shared their lived experiences and perspectives to ensure that the knowledge we co-create addresses the concerns of patients and the public. Creating the largest network of collaboration across Canadian Emergency Departments would not have been feasible without the tireless efforts of Emergency Department Chiefs, and research coordinators and research assistants at participating sites. Finally, our most humble and sincere gratitude to all our colleagues in medicine, nursing, and the allied health professions who have been on the front lines of this pandemic from day one staffing our ambulances, Emergency Departments, ICUs, and hospitals bravely facing the risks of COVID-19 to look after our fellow citizens and after one another. We dedicate this network to you. (Supplementary Table)

## Data Sharing

For investigators who wish to access Canadian COVID-19 Emergency Rapid Response Network data, proposals may be submitted to the network for review and approval by the network’s peer-review publication committee, the data access and management committee and the executive committee, as per the network’s governance. Information regarding submitting proposals and accessing data may be found at https://canadiancovid19ednetwork.org/.

## Funding Acknowledgement

The network is funded by the Canadian Institutes of Health Research (447679), BC Academic Health Science Network Society, BioTalent Canada, Genome BC (COV024; VAC007), Ontario Ministry of Colleges and Universities (C-655-2129), the Saskatchewan Health Research Foundation (5357) and the Fondation CHU de Québec (Octroi #4007).

## Appendices

**Appendix Table 1.**
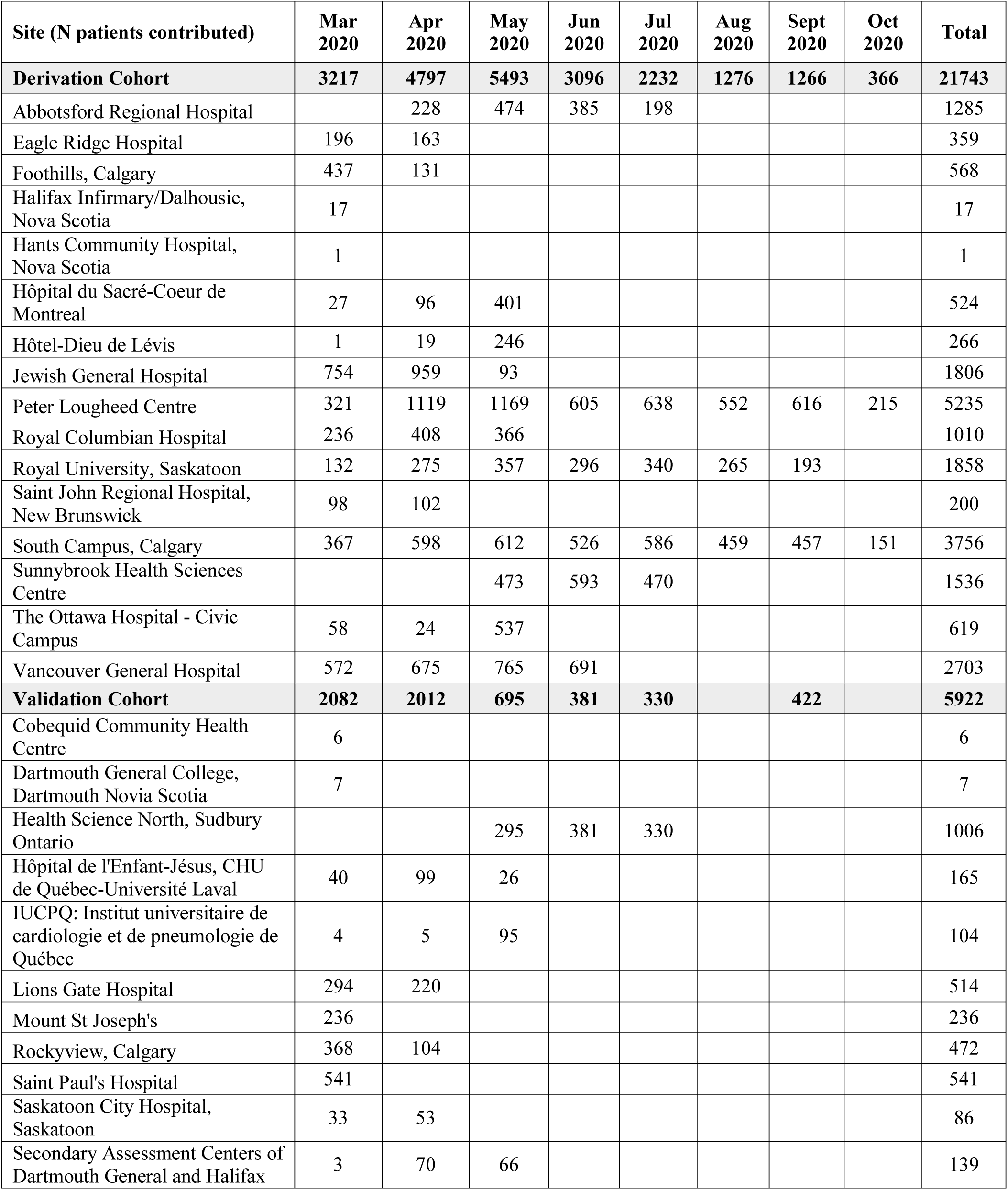
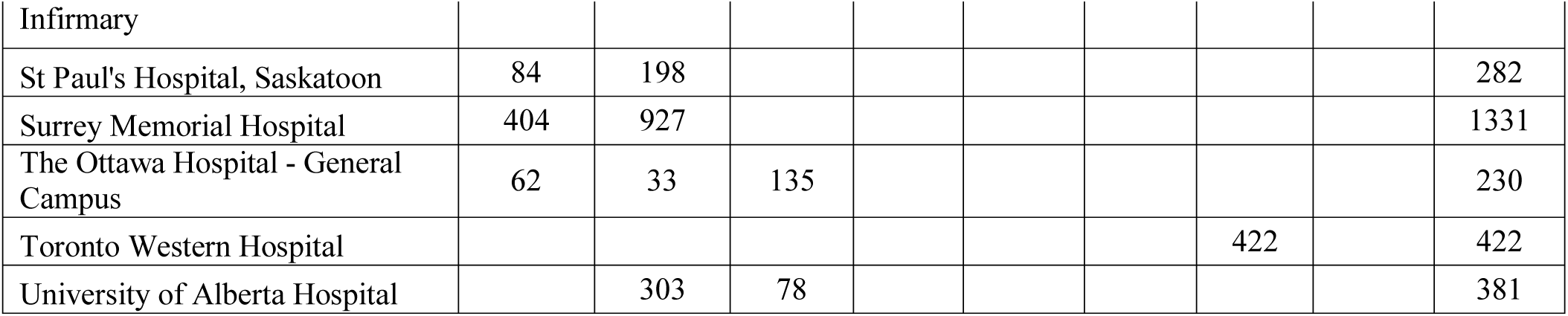
Patients enrolled by CCEDRRN site and time periods for data collection

**Appendix Table 2.** Candidate variables for entry into regression model

| Variable | Definition | N (%)<br>Missing |
| --- | --- | --- |
| **Demographics** |  |  |
| Age | Age in years | 0 (0) |
| Sex | Male, Female, Other | 0 (0) |
| Arrival from | Home + other (not clearly documented) | 0 (0) |
|  | Single room + no fixed address + shelter | 0 (0) |
|  | Institutional living: long-term care/rehab + correctional | 0 (0) |
|  | Inter-hospital transfer | 0 (0) |
| **Infection risk** |  |  |
| Travel risk | Travel from country with known cases within 14 days | 0 (0) |
| Institutional exposure | Possible exposure in institutional setting (e.g., Long-term care, prison) | 0 (0) |
| Healthcare worker | Healthcare worker/Microbiology lab employee | 0 (0) |
| Household/caregiver contact | Household contact /caregiver of known positive case | 0 (0) |
| No documented risk | Documented absence of risk factors | 0 (0) |
| **Emergency department variables** |  |  |
| ED arrival mode |  | 2 (0) |
| Ambulance: | arrived by ambulance |  |
| Self/police | self-transported or transported to ED by police |  |
| Arrival heart rate | beats/minute | 452 (2.1) |
| Arrival respiratory rate | breaths/minute | 732 (3.4) |
| Arrival oxygen saturation | % | 517 (2.4) |
| Lowest recorded oxygen saturation in ED | % | 445 (2.0) |
| Fever |  | 847 (3.9) |
| Temperature <36.0 | Temperature <36.0C AND no self-reported fever |  |
| Temperature 36.0-37.4 | Temperature 36.0-37.4C AND no self-reported fever |  |
| Temperature ≥37.5 or fever | Temperature ≥37.5 OR self-reported fever |  |
| Respiratory distress | Increased work of breathing documented by treating clinician | 1(0) |
| Supplemental oxygen delivered in the ED | Yes/No | 0 (0) |
| **COVID symptoms** |  |  |
| Abdominal pain | Patient-reported symptom as documented by treating clinician | 0 (0) |
| Altered consciousness/confusion | Patient-reported symptom as documented by treating clinician | 0 (0) |
| Bleeding (hemorrhage) | Patient-reported symptom as documented by treating clinician | 0 (0) |
| Chest pain (includes discomfort or tightness) | Patient-reported symptom as documented by treating clinician | 0 (0) |
| Chills | Patient-reported symptom as documented by treating clinician | 0 (0) |
| Conjunctivitis | Patient-reported symptom as documented by treating clinician | 0 (0) |
| Cough | Patient-reported symptom as documented by treating clinician | 0 (0) |
| Diarrhea | Patient-reported symptom as documented by treating clinician | 0 (0) |
| Dizziness/Vertigo | Patient-reported symptom as documented by treating clinician | 0 (0) |
| Dysgeusia/anosmia | Patient-reported symptom as documented by treating clinician | 0 (0) |
| Ear pain | Patient-reported symptom as documented by treating clinician | 0 (0) |
| Fatigue/malaise | Patient-reported symptom as documented by treating clinician | 0 (0) |
| Headache | Patient-reported symptom as documented by treating clinician | 0 (0) |
| Hemoptysis (bloody sputum) | Patient-reported symptom as documented by treating clinician | 0 (0) |
| Joint pain (arthralgia) | Patient-reported symptom as documented by treating clinician | 0 (0) |
| Lymphadenopathy | Patient-reported symptom as documented by treating clinician | 0 (0) |
| Muscle aches (myalgia) | Patient-reported symptom as documented by treating clinician | 0 (0) |
| Nausea/vomiting | Patient-reported symptom as documented by treating clinician | 0 (0) |
| No reported symptoms | Patient-reported symptom as documented by treating clinician | 0 (0) |
| Runny nose (rhinorrhea) | Patient-reported symptom as documented by treating clinician | 0 (0) |
| Seizures | Patient-reported symptom as documented by treating clinician | 0 (0) |
| Shortness of breath (dyspnea) | Patient-reported symptom as documented by treating clinician | 0 (0) |
| Skin rash | Patient-reported symptom as documented by treating clinician | 0 (0) |
| Sore throat | Patient-reported symptom as documented by treating clinician | 0 (0) |
| Sputum production | Patient-reported symptom as documented by treating clinician | 0 (0) |
| Wheezing | Patient-reported symptom as documented by treating clinician | 0 (0) |
| Current tobacco user | Documented current tobacco use | 6 (0) |
| Current illicit user | Documented methamphetamine, opioid or other illicit drug use | 6 (0) |
| **7-day average incident COVID-19 cases** | Daily reported incidence of new cases in health region, averaged over the seven days preceding hospital arrival. Reported in units of new cases/100,000 population | 32 (0.1) |

**Appendix Table 3.**
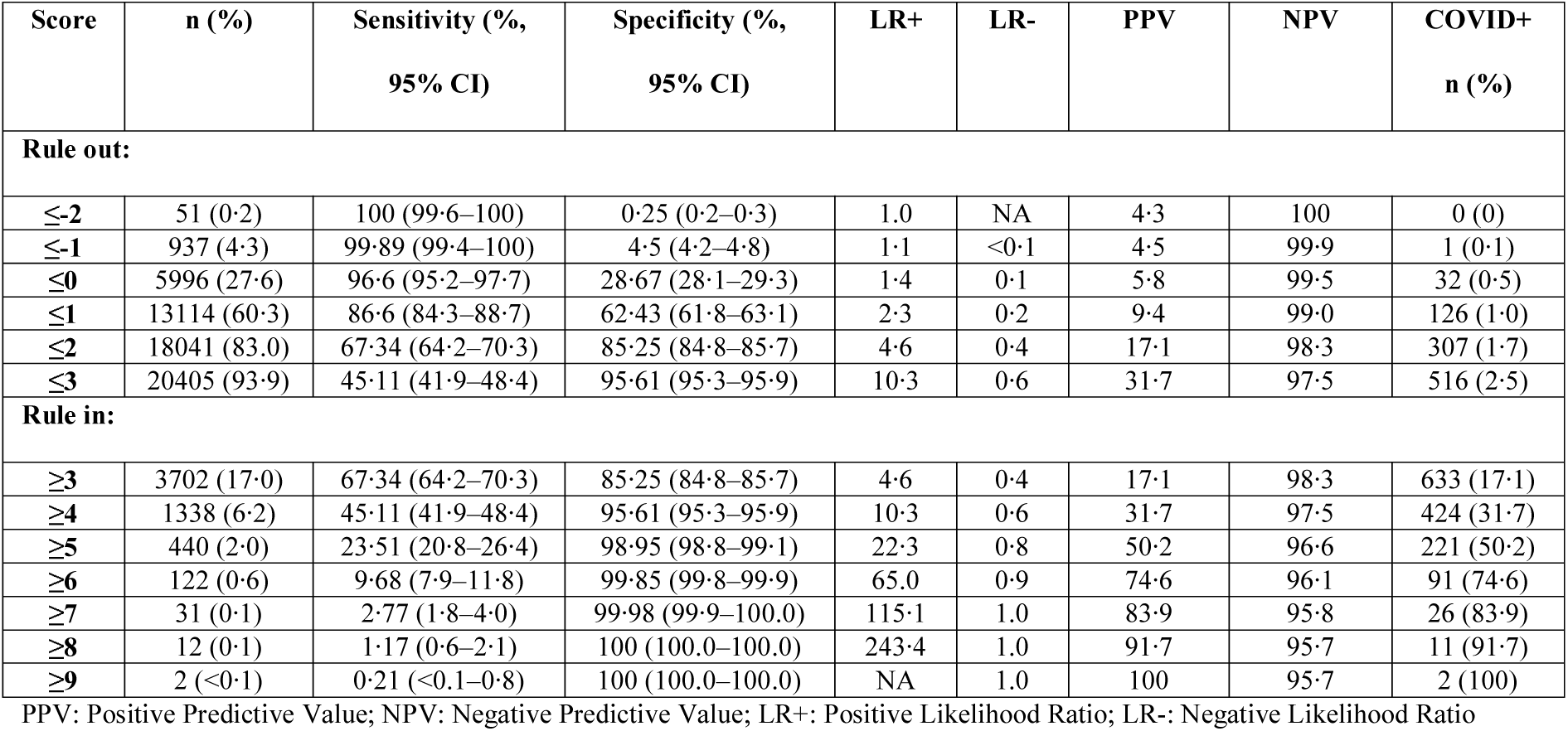
Performance metrics for the CCEDRRN COVID-19 Infection Score for ruling in or ruling out SARS-CoV-2 infection at different score cut-off values in the derivation cohort

**Appendix Table 4.** Performance metrics for CCEDRRN COVID Infection Score for ruling in or ruling out SARS-CoV-2 infection at different score cut-off values in the combined cohort

| Score cutoff | n (%) | Sensitivity (95% CI) | Specificity (95% CI) | PPV | NPV | LR+ | LR- | COVID+ n (%) |
| --- | --- | --- | --- | --- | --- | --- | --- | --- |
| **Rule out:** |  |  |  |  |  |  |  |  |
| ≤-2 | 70 (0.3) | 100 (99.7,100) | 0.3 (0.2,0.3) | 4.2 | 100 | 1 | 0 | 0 (0) |
| ≤-1 | 1257 (4.5) | 99.8 (99.4,100) | 4.7 (4.5,5) | 4.4 | 99.8 | 1.1 | <0.1 | 2 (0.2) |
| ≤0 | 7872 (28.5) | 96.1 (94.8,97.1) | 29.5 (29.0,30.1) | 5.7 | 99.4 | 1.4 | 0.1 | 46 (0.6) |
| ≤1 | 16962 (61.3) | 85 (82.8,87.0) | 63.4 (62.8,63.9) | 9.3 | 99.0 | 2.3 | 0.2 | 175 (1.0) |
| ≤2 | 23243 (84.0) | 64.8 (62.0,67.5) | 86.2 (85.7,86.6) | 17.1 | 98.2 | 4.7 | 0.4 | 411 (1.8) |
| ≤3 | 26169 (94.6) | 40.3 (37.4,43.2) | 96.1 (95.9,96.4) | 31.4 | 97.3 | 10.4 | 0.6 | 697 (2.7) |
| **Rule in:** |  |  |  |  |  |  |  |  |
| ≥3 | 4422 (16.0) | 64.8 (62.0,67.5) | 86.2 (85.7,86.6) | 17.1 | 98.2 | 4.7 | 0.4 | 756 (17.1) |
| ≥4 | 1496 (5.4) | 40.3 (37.4,43.2) | 96.1 (95.9,96.4) | 31.4 | 97.3 | 10.4 | 0.6 | 470 (31.4) |
| ≥5 | 476 (1.7) | 20.2 (18.0,22.6) | 99.1 (99.0,99.2) | 49.6 | 96.6 | 22.3 | 0.8 | 236 (49.6) |
| ≥6 | 128 (0.5) | 8 (6.5,9.7) | 99.9 (99.8,99.9) | 72.7 | 96.1 | 60.3 | 0.9 | 93 (72.7) |
| ≥7 | 32 (0.1) | 2.2 (1.5,3.2) | 100 (100,100) | 81.2 | 95.9 | 98.4 | 1.0 | 26 (81.3) |
| ≥8 | 12 (<0.1) | 0.9 (0.5,1.7) | 100 (100,100) | 91.7 | 95.8 | 249.8 | 1.0 | 11 (91.7) |
| ≥9 | 2 (<0.1) | 0.2 (0,0.6) | 100 (100,100) | 100 | 95.8 | Inf | 1.0 | 2 (100) |

TP: True positives; FP: False positives; TN: True negatives; FN: False negatives; PPV: Positive Predictive Value; NPV: Negative Predictive Value; LR+: Positive likelihood ratio; LR-: Negative likelihood ratio

**Appendix Table 5.**
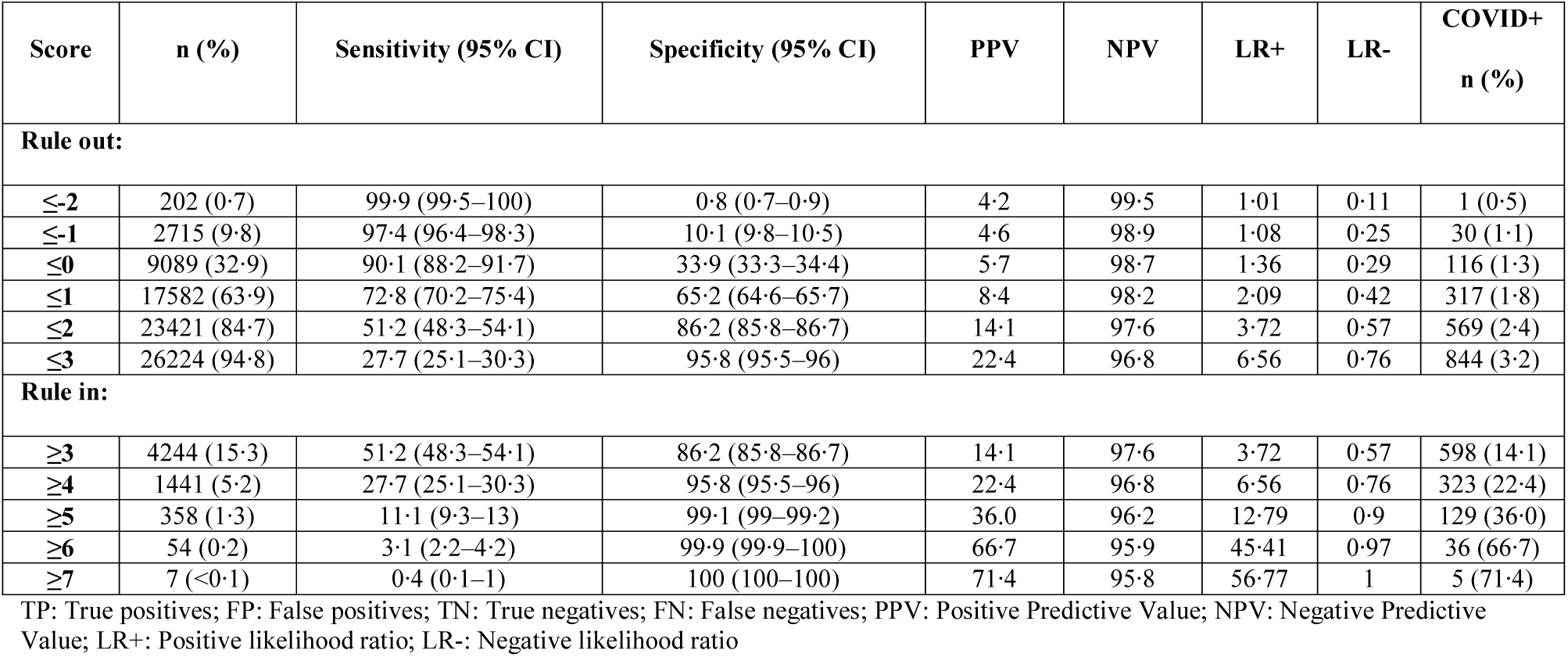
Performance metrics for the CORC score (race and ethnicity variables removed) for ruling in or ruling out SARS-CoV-2 infection at different score cut-off values in the combined cohort

| Score | n (%) | Sensitivity (95% CI) | Specificity (95% CI) | PPV | NPV | LR+ | LR- | COVID+<br>n (%) |
| --- | --- | --- | --- | --- | --- | --- | --- | --- |
| **Rule out:** |  |  |  |  |  |  |  |  |
| ≤-2 | 202 (0·7) | 99·9 (99·5–100) | 0·8 (0·7–0·9) | 4·2 | 99·5 | 1·01 | 0·11 | 1 (0·5) |
| ≤-1 | 2715 (9·8) | 97·4 (96·4–98·3) | 10·1 (9·8–10·5) | 4·6 | 98·9 | 1·08 | 0·25 | 30 (1·1) |
| ≤0 | 9089 (32·9) | 90·1 (88·2–91·7) | 33·9 (33·3–34·4) | 5·7 | 98·7 | 1·36 | 0·29 | 116 (1·3) |
| ≤1 | 17582 (63·9) | 72·8 (70·2–75·4) | 65·2 (64·6–65·7) | 8·4 | 98·2 | 2·09 | 0·42 | 317 (1·8) |
| ≤2 | 23421 (84·7) | 51·2 (48·3–54·1) | 86·2 (85·8–86·7) | 14·1 | 97·6 | 3·72 | 0·57 | 569 (2·4) |
| ≤3 | 26224 (94·8) | 27·7 (25·1–30·3) | 95·8 (95·5–96) | 22·4 | 96·8 | 6·56 | 0·76 | 844 (3·2) |
| **Rule in:** |  |  |  |  |  |  |  |  |
| ≥3 | 4244 (15·3) | 51·2 (48·3–54·1) | 86·2 (85·8–86·7) | 14·1 | 97·6 | 3·72 | 0·57 | 598 (14·1) |
| ≥4 | 1441 (5·2) | 27·7 (25·1–30·3) | 95·8 (95·5–96) | 22·4 | 96·8 | 6·56 | 0·76 | 323 (22·4) |
| ≥5 | 358 (1·3) | 11·1 (9·3–13) | 99·1 (99–99·2) | 36·0 | 96·2 | 12·79 | 0·9 | 129 (36·0) |
| ≥6 | 54 (0·2) | 3·1 (2·2–4·2) | 99·9 (99·9–100) | 66·7 | 95·9 | 45·41 | 0·97 | 36 (66·7) |
| ≥7 | 7 (<0·1) | 0·4 (0·1–1) | 100 (100–100) | 71·4 | 95·8 | 56·77 | 1 | 5 (71·4) |

TP: True positives; FP: False positives; TN: True negatives; FN: False negatives; PPV: Positive Predictive Value; NPV: Negative Predictive Value; LR+: Positive likelihood ratio; LR-: Negative likelihood ratio

**Appendix Table 6.** Net Reclassification Improvement of the CCEDRRN COVID-19 Infection Score compared to the CORC Score (race and ethnicity variables removed)

| **Primary Outcome : Covid Positive** |  |  |  |  |  |
| --- | --- | --- | --- | --- | --- |
| **CORC risk category** |  | **CCIS risk category** |  |  | **Total** |
|  |  | **Low** | **Medium** | **High** |  |
|  | **Low** | 12 | 18 | 0 | 30 |
|  | **Medium** | 34 | 539 | 241 | 814 |
|  | **High** | 0 | 94 | 229 | 323 |
|  | **Total** | 46 | 651 | 470 | 1167 |
| **Primary Outcome: Covid Negative** |  |  |  |  |  |
| **CORC risk category** |  | **CCIS risk category** |  |  | **Total** |
|  |  | **Low** | **Medium** | **High** |  |
|  | **Low** | 1593 | 1092 | 0 | 2685 |
|  | **Medium** | 6233 | 15756 | 706 | 22695 |
|  | **High** | 0 | 798 | 320 | 1118 |
|  | **Total** | 7826 | 17646 | 1026 | 26498 |

| **<u>COVID Positive</u>** |  | **<u>COVID Negative</u>** |  |
| --- | --- | --- | --- |
| Number of outcomes | 1167 | Number of outcomes | 26498 |
| Correct reclassification | 259 | Correct reclassification | 7031 |
| Incorrect reclassification | 128 | Incorrect reclassification | 1798 |
| Net reclassification | 131 | Net reclassification | 5233 |
| Net reclassification improvement (Event) | 0.112 | Net reclassification improvement (Non-event) | 0.197 |
| Total net reclassification improvement |  |  | 0.310 |

**Appendix Figure 1.**
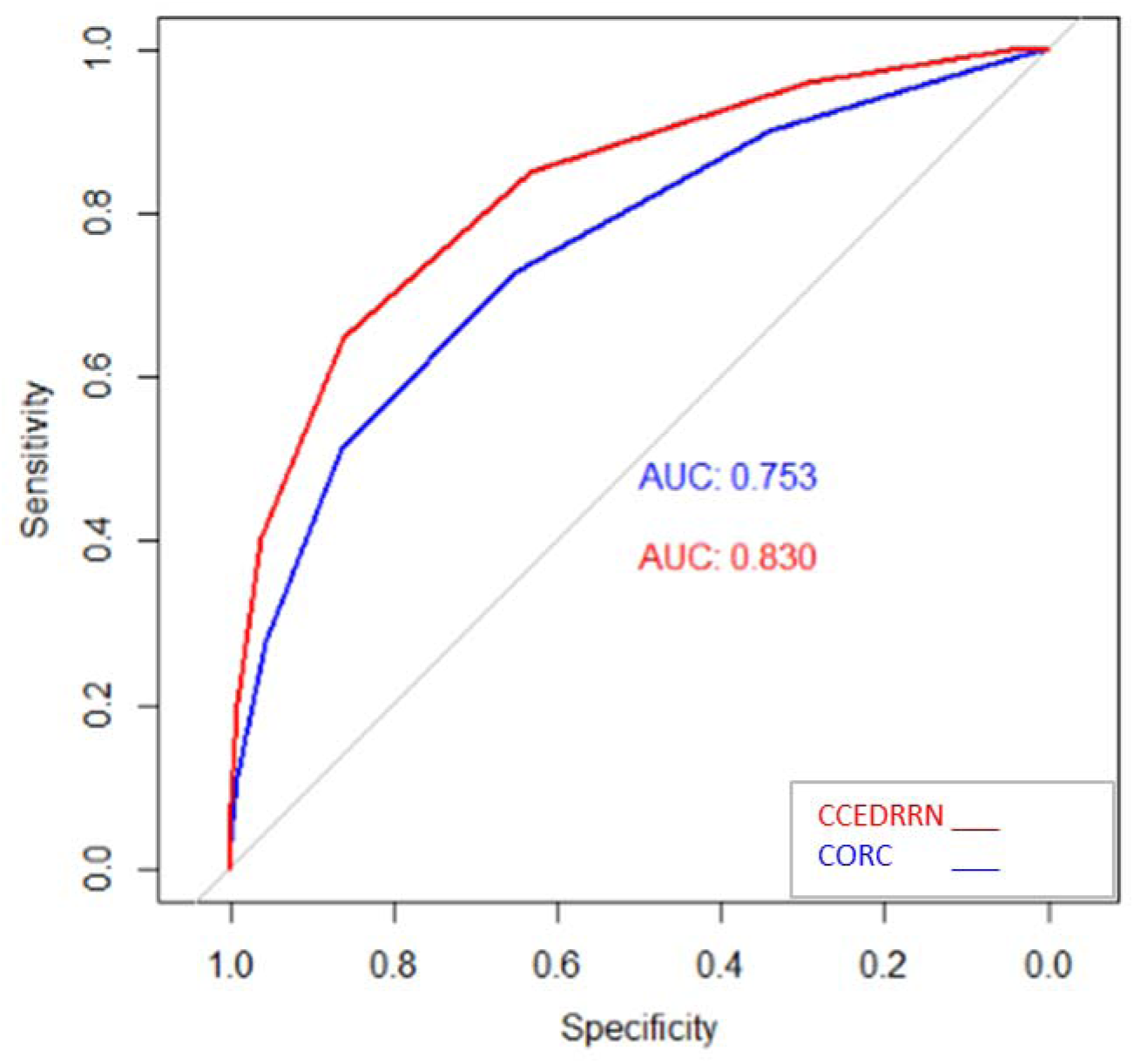
Receiver operating characteristic curves for the CCEDRRN COVID Infection Score (CCIS) and the CORC score (race and ethnicity variables removed) in the combined study cohort

## Research in context

### Evidence before this study

We searched the Medline, Embase and Cochrane Central Library for studies describing the derivation and/or validation of risk scores that estimate the probability of a positive SARS-CoV-2 nucleic acid test result in emergency department patients with suspected coronavirus-19 (COVID-19) disease using simple clinical variables. Search terms included (COVID-19 OR SARS-CoV-2 OR coronavirus) and (Emergency Services, Hospital OR Accident OR Accident and emergency OR emergency medical services), and limited to the pandemic period (November 1, 2019 o December 1, 2020.). We also identified risk scores from a living systematic review that was most recently updated on February 1, 2021. We identified 10 risk scores for predicting SARS-CoV-2 infection in emergency department patients. All but one risk score included laboratory or diagnostic imaging results in addition to clinical variables. Several employed machine learning approaches that would require an advanced electronic medical record for implementation. The only risk prediction tool limited to clinical variables was derived in a population with a high proportion of SARS-CoV-2-positive patients, and is thus vulnerable to selection bias. This risk score also included three race or ethnicity variables, which may limit its generalizability is limited. outside of the population in which it was developed.

### Added value of this study

Using data from pandemic waves 1 and 2 in a large, geographically distributed multicenter emergency department research network in Canada (https://canadiancovid19ednetwork.org/), we have derived and validated a user-friendly 10-item risk prediction tool that uses variables available at the time of a patient’s initial presentation. Our tool accurately excludes COVID-19 infection in one-third of patients and accurately rules in COVID-19 infection in high-risk patients. This risk score includes a measure local COVID-19 incidence to account for local SARS-CoV-2 transmission patterns. This risk score is generalizable across geographic settings and does not require diagnostic tests or advanced electronic decision support for implementation.

### Implications of all the available evidence

In a diverse set of hospitals, we developed and externally validated a risk score to quantify the probability of SARS-CoV-2 infection in emergency department patients. This score can guide isolation practices from the time of a patient’s hospital arrival. Patients classified as low-risk need not be tested, which is advantageous is low in settings where resources are limited. Patients classified as high-risk can be prioritized for rapid testing, isolation and/or early initiation of empiric therapy prior to the availability of COVID-19 test results.

