## Supplementary material for "Development and validation of a clinical risk score to predict SARS-CoV-2 infection in emergency department patients: The CCEDRRN COVID-19 Infection Score (CCIS)": Acknowledgement Table (from another version of this preprint: version 1, 2021-07-19)

### Supplementary Table: Contributors to the Canadian COVID-19 Emergency Department Rapid Response Network

#### 1. Purpose

This supplementary table provides details of the support staff at each of the participating institutions in the Canadian COVID-19 Emergency Department Rapid Response Network. This supplementary document should be attached to each peer-reviewed manuscript after the methods manuscript (M1). The purpose is to ensure research staffs and lead coordinators are appropriately recognized for their contributions to the network.

#### 2. List of Support Staff

Table 1. Network coordinating centre staff at the University of British Columbia

| Name | Roles | Contributions |
|---|---|---|
| Gelareh Ghaderi | Data analyst | Data processing and analysis for manuscripts. |
| Jeffrey Hau | Data manager | REDCap, data processing and analysis for manuscripts. |
| Vi Ho | National coordinator | Coordinate with provincial coordinators and training/onboarding of research assistants. |
| Joe Larkin | Project manager | Project management. |
| Fiona O'Sullivan | Data analyst | Data processing and analysis for manuscripts. |
| Serena Small | Research coordinator | Ethics & privacy reviews, data management plan, privacy impact assessment, and qualitative analyses |
| Amber Cragg | Research manager | Data and manuscript management |
| Wei Zhao | Data analyst | Data processing and analysis for manuscripts. |
| Vicky Wu | Data analyst | Data processing and analysis for manuscripts. |
| Elnaz Bodaghkhani | Research associate | Data and manuscript management |

Table 2. Provincial Coordinators

| Name | Province | Institutional affiliation | Contributions to CCEDRRN |
|---|---|---|---|
| Corinne DeMone | NS | Dalhousie University, Halifax, Nova Scotia | Research ethics board submission, manages research assistants, data cleaning and quality. |
| Jacqueline Fraser | NB | Dalhousie University, St. John New Brunswick | Site coordinator as well as research assistant. |
| Veronique Gélinas | QC | Centre intégré de santé et de services sociaux de Chaudière-Appalaches (Hôtel-Dieu de Lévis site), Lévis | Provincial research coordinator, translation of research material to French, ethics management. |

| | | | |
|---|---|---|---|
| Connie Taylor | ON | Queen's University, Kingston | Coordination of research assistants in Ontario, maintenance of REB applications for the province |
| Kate Mackenzie | MB | Health Sciences Centre, Winnipeg | Lead RA for the province |
| Aimee Goss | SK | University of Saskatchewan, Saskatoon | Screens records in Saskatoon, data/extraction and entry, coordinates research assistants. |
| Hina Walia | AB | University of Calgary, Calgary | Provincial coordinator lead for Alberta, oversight of all Alberta sites. |
| Rajan Bola | BC | University of British Columbia, Vancouver | Provincial coordinator lead for BC, oversight of all BC sites. |

Table 3. Institutional research assistant (RA) leads
Institutional RA leads are responsible for data extraction and integrity, communication with provincial leads.

| **Name** | **Province** | **Institutional affiliation(s)** |
|---|---|---|
| Corinne DeMone | NS | Dartmouth General Hospital, Cobequid Community Health Centre, Hants Community Hospital<br>Secondary Assessment Centers of the Dartmouth General Hospital, and Halifax Infirmary, Halifax |
| Jacqueline Fraser | NB | Saint John Regional Hospital, Saint John |
| Alexandra Nadeau | QC | CHU de Québec Université Laval, Quebec City |
| Audrey Nolet | QC | Centre intégré de santé et de services sociaux de Chaudière-Appalaches (Hôtel-Dieu de Lévis site), Lévis |
| Xiaoqing Xue | QC | Jewish General Hospital, Montréal |
| David Iannuzzi | QC | McGill University Health Center, Montréal |
| Chantal Lanthier | QC | Hôpital du Sacré-Cœur de Montréal, Montréal |
| Konika Nirmalanathan | ON | University Health Network, Toronto |
| Vlad Latiu | ON | Kingston General Hospital, Hotel Dieu Hospital, Kingston |
| Joanna Yeung | ON | Sunnybrook Health Sciences Center, Toronto |
| Natasha Clayton | ON | Hamilton General Hospital, Juravinski Hospital, Hamilton |
| Tom Chen | ON | London Health Sciences Centre, London |
| Jenna Nichols | ON | Health Sciences North, Sudbury |
| Kate Mackenzie | MB | Health Sciences Centre, Winnipeg |
| Aimee Goss | SK | St. Paul's Hospital, Royal University Hospital, Saskatoon City Hospital, Saskatoon |
| Stacy Ruddell | AB | Foothills Medical Centre, Peter Lougheed Centre, Rockyview General Hospital, South Health Campus, Calgary |
| Natalie Runham | AB | University of Alberta Hospital, Edmonton |

| Name | Province | Institutional affiliation(s) |
|---|---|---|
| Karlin Su | AB | Royal Alexandra Hospital/Northeast Community Health Center, Edmonton |
| Josie Kanu | BC | St. Paul's Hospital, Mount Saint Joseph, Vancouver |
| Bernice Huynh | BC | Abbotsford Regional Hospital and Cancer Center, Abbotsford |
| Amanda Swirhun | BC | Royal Columbian Hospital, New Westminster |
| Tracy Taylor | BC | Eagle Ridge Hospital and Health Care Centre, Port Moody |
| Mai Hayashi | BC | Royal Inland Hospital, Kamloops |
| Mackenzie Cheyne | BC | Kelowna General Hospital, Kelowna |
| Sarim Asim | BC | Surrey Memorial Hospital, Surrey |
| Katherine Lam | BC | Vancouver General Hospital, Vancouver |
| Kelsey Compagna | BC | Lions Gate Hospital, Vancouver |

Table 4. Contributing Study Sites and Investigators

| Lead Investigator | Contributing Site / Code | Member Investigators |
|---|---|---|
| **Maritime** | | |
| Patrick Fok | | |
| **Nova Scotia** | | |
| Hana Wiemer | Halifax Infirmary/ 902 | Patrick Fok |
| | Dartmouth General Hospital/ 903 | Hana Wiemer |
| | Hants Community Hospital/ 904 | Samuel Campbell |
| | Cobequid Community Health Centre/ 905 | Kory Arsenault |
| | Secondary Assessment Centers of Dartmouth General and Halifax Infirmary/ 908 | Tara Dahn |
| **New Brunswick** | | |
| Kavish Chandra | Saint John Regional Hospital/ 901 | Kavish Chandra |
| **Quebec** | | |
| Patrick Archambault | Hotel-Dieu de Lévis/ 701 | Patrick Archambault |
| | Jewish General Hospital/ 702 | Joel Turner |
| | Centre Hospitalier de l'Université Laval (CHU de Québec)/ 703 | Éric Mercier |
| | L'hôpital Royal Victoria - Royal Victoria Hospital/ 705 | Greg Clark |
| | Hôpital de l'Enfant-Jésus,CHU de Québec/ 706 | Éric Mercier |
| | Hôpital du Saint-Sacrement, CHU de Québec/ 707 | Éric Mercier |
| | Hôpital Saint-François d'Assise, CHU de Québec/ 708 | Éric Mercier |
| | Hôtel-Dieu de Québec,CHU de Québec/ 709 | Éric Mercier |

| | | |
|---|---|---|
| | IUCPQ: Institut universitaire de cardiologie et de pneumologie de Québec/ 710 | Sébastien Robert |
| | Hôpital du Sacré-Coeur de Montreal/ 711 | Raoul Daoust |
| **Ontario** | | |
| Laurie Morrison & Steven Brooks | Sunnybrook/ 401 | Ivy Cheng |
| | The Ottawa Hospital - Civic Campus/ 403 | Krishan Yadav |
| | The Ottawa Hospital - General Campus/ 404 | Krishan Yadav |
| | Kingston/Queens/ 406 | Steven Brooks |
| | Hamilton General Hospital/ 407 | Michelle Welsford |
| | Health Science North, Sudbury Ontario/ 408 | Rob Ohle |
| | University Hospital – LHSC/ 409 | Justin Yan |
| | North York General Hospital, Toronto/ 410 | Rohit Mohindra |
| | Victoria Hospital – LHSC/ 412 | Justin Yan |
| | Toronto Western Hospital/ 414 | Megan Landes |
| **Manitoba** | | |
| Tomislav Jelic | Health Sciences Centre/ 307 | Tomislav Jelic |
| **Saskatchewan** | | |
| Phil Davis | Pasqua Hospital, Regina/ 301 | Ankit Kapur |
| | Regina General Hospital, Regina/ 302 | Ankit Kapur |
| | St Paul's Hospital, Saskatoon/ 303 | Phil Davis |
| | Royal University, Saskatoon/ 304 | Phil Davis |
| | Saskatoon City Hospital, Saskatoon/ 305 | Phil Davis |
| **Alberta** | | |
| Andrew McRae | University of Alberta Hospital, Edmonton/ 201 | Brian Rowe |
| | Foothills, Calgary/ 202 | Katie Lin |
| | Rockyview, Calgary/ 203 | Andrew McRae |
| | Peter Lougheed Centre/ 204 | Andrew McRae |
| | South Campus, Calgary/ 205 | Stephanie VandenBerg |
| | Northeast Community Health Centre, Edmonton/ 206 | Jake Hayward, Jaspreet Khangura |
| | Royal Alexandra Hospital, Edmonton/ 306 | Jake Hayward, Jaspreet Khangura |
| **British Columbia** | | |
| Corinne Hohl | Vancouver General Hospital/ 101 | Daniel Ting |
| | Lions Gate Hospital/ 102 | Maja Stachura |
| | Saint Paul's Hospital/ 103 | Frank Scheuermeyer |

| | |
|---|---|
| Mount St Joseph's/ 104 | Frank Scheuermeyer |
| Surrey Memorial Hospital/ 105 | Balijeet Braar/ Craig Murray |
| Royal Columbian Hospital/ 106 | John Taylor |
| Abbotsford Regional Hospital/ 107 | Ian Martin |
| Eagle Ridge Hospital/ 108 | Sean Wormsbecker |
| Victoria General Hospital/ 109 | Matt Bouchard |
| Royal Jubilee Hospital/ 110 | Matt Bouchard |
| Nanaimo General Hospital/ 111 | Matt Bouchard |
| Royal Inland Hospital/ 112 | Ian Martin |
| Kelowna General / Hospital/ 115 | Lee Graham |

It was not possible for us to recruit Members from Newfoundland and Labrador, Northwest Territories, Nunavut, Prince Edward Island and Yukon at the time of the inception of the registry.
